# Longitudinal diffusion and volumetric kinetics of head and neck cancer magnetic resonance on a 1.5T MR-Linear accelerator hybrid system: A prospective R-IDEAL Stage 2a imaging biomarker characterization/ pre-qualification study

**DOI:** 10.1101/2023.05.04.23289527

**Authors:** Joint Head and Neck Radiation Therapy-MRI Development Cooperative, MR-Linac Consortium Head and Neck Tumor Site Group, Dina M El-Habashy, Kareem A Wahid, Renjie He, Brigid McDonald, Jillian Rigert, Samuel J. Mulder, Tze Yee Lim, Xin Wang, Jinzhong Yang, Yao Ding, Mohamed A Naser, Sweet Ping Ng, Houda Bahig, Travis C Salzillo, Kathryn E Preston, Moamen Abobakr, Mohamed A Shehata, Enas A Elkhouly, Hagar A Alagizy, Amira H Hegazy, Mustefa Mohammadseid, Chris Terhaard, Marielle Philippens, David I. Rosenthal, Jihong Wang, Stephen Y. Lai, Alex Dresner, John C. Christodouleas, Abdallah Sherif Radwan Mohamed, Clifton D Fuller

## Abstract

**Objectives:** We aim to characterize the serial quantitative apparent diffusion coefficient (ADC) changes of the target disease volume using diffusion-weighted imaging (DWI) acquired weekly during radiation therapy (RT) on a 1.5T MR-Linac and correlate these changes with tumor response and oncologic outcomes for head and neck squamous cell carcinoma (HNSCC) patients as part of a programmatic R-IDEAL biomarker characterization effort.

**Methods:** Thirty patients with pathologically confirmed HNSCC who received curative-intent RT at the University of Texas MD Anderson Cancer Center, were included in this prospective study. Baseline and weekly Magnetic resonance imaging (MRI) (weeks 1-6) were obtained, and various ADC parameters (mean, 5^th^, 10^th^, 20^th^, 30^th^, 40^th^, 50^th^, 60^th^, 70^th^, 80^th^, 90^th^ and 95^th^ percentile) were extracted from the target regions of interest (ROIs). Baseline and weekly ADC parameters were correlated with response during RT, loco-regional control, and the development of recurrence using the Mann-Whitney U test. The Wilcoxon signed-rank test was used to compare the weekly ADC versus baseline values. Weekly volumetric changes (Δvolume) for each ROI were correlated with ΔADC using Spearman’s Rho test. Recursive partitioning analysis (RPA) was performed to identify the optimal ΔADC threshold associated with different oncologic outcomes.

**Results:** There was an overall significant rise in all ADC parameters during different time points of RT compared to baseline values for both gross primary disease volume (GTV-P) and gross nodal disease volumes (GTV-N). The increased ADC values for GTV-P were statistically significant only for primary tumors achieving complete remission (CR) during RT. RPA identified GTV-P ΔADC 5^th^ percentile >13% at the 3^rd^ week of RT as the most significant parameter associated with CR for primary tumor during RT (p <0.001). Baseline ADC parameters for GTV-P and GTV-N didn’t significantly correlate with response to RT or other oncologic outcomes. There was a significant decrease in residual volume of both GTV-P & GTV-N throughout the course of RT. Additionally, a significant negative correlation between mean ΔADC and Δvolume for GTV-P at the 3^rd^ and 4^th^ week of RT was detected (r = -0.39, p = 0.044 & r = -0.45, p = 0.019, respectively).

**Conclusion:** Assessment of ADC kinetics at regular intervals throughout RT seems to be correlated with RT response. Further studies with larger cohorts and multi-institutional data are needed for validation of ΔADC as a model for prediction of response to RT.

## INTRODUCTION

Head and neck squamous cell carcinoma (HNSCC) are the seventh most common malignancy worldwide [1], for which radiation therapy (RT) with or without systemic therapy, is considered the mainstay treatment [2]. Complementary RT techniques have enabled more conformal dose distributions allowing maximum dose delivery to tumor volume while sparing the surrounding normal tissue below standard threshold doses [3]. Despite remarkable progress in RT techniques, treatment outcome is still unsatisfactory, especially for cases of advanced stage HNSCC [4].

The ability to use a readily available, and easily interpretable biomarker to characterize tumors as resistant or sensitive, and adapt treatment accordingly remains an unmet need [5]. To date magnetic resonance imaging (MRI) has been shown to be able to predict treatment response and oncologic outcomes [6]. While anatomical MRI has been used for accurate tumor definition and delineation because of its superior soft tissue contrast compared to CT, functional MRI can assess physiologic changes that may be undetected by the naked eye [7].

MRI linear accelerators (MR-Linacs) are innovative RT devices in which a linear accelerator is integrated with an on-board MRI scanner to enable MR-guided adaptive RT [8]. In addition to standard anatomical MRI sequences, functional MRI sequences such as diffusion-weighted imaging (DWI) can be acquired on these devices [9]. Therefore, acquisition of daily quantitative images, which is not feasible using standard diagnostic MRI protocols, is now possible using the MR-Linac. These quantitative imaging techniques provide information not only about tissue characteristics but also about the potential tumoral changes throughout the course of treatment [10].

Prior to and during implementation of innovative radiation technology such as the MR-Linac, it is important to have a standard method to evaluate benefits, risks, and outcomes. In 2009, the Balliol Consortium created the IDEAL framework, which includes five stages for assessing surgical innovations: Idea, Development, Exploration, Assessment, and Long-term evaluation [11]. The R-IDEAL framework is a similar stage-based process for developing and evaluating radiotherapy innovations [12]. The MR-Linac consortium, an international collaboration towards advancing MR-Linac technology, has adopted the R-IDEAL framework to ensure a coordinated and evidence-based introduction of the MR-Linac to clinics [13].

DWI is a specific MRI technique with potential utility for evaluating and predicting tumor response to treatment [14]. This technique depends on the Brownian motion of water molecules within tissues, which depends on cellular density, cell wall integrity, and vascularity. The diffusion through each voxel is quantified through the apparent diffusion coefficient (ADC), a quantitative imaging biomarker [15]. ADC was found to be correlated with tumor response to treatment in different types of malignancies [16-18]. Additionally, several studies have elaborated the role of ADC as a potential biomarker in head and neck cancer [19-23].

Our group has demonstrated that mid-RT ADC changes can identify patients who are more likely to achieve complete remission for their primary tumor by the end of RT for human papillomavirus (HPV) positive oropharyngeal carcinoma [24]. In the same context, another study completed by our group has reported that the primary tumors’ change in ADC (ΔADC) at mid-RT is a robust predictor of oncologic outcomes in patients with head and neck cancer [25]. Moreover, a recent study has shown that DWI-guided dose painting had a better disease-free survival compared to conventional intensity-modulated RT (IMRT) in patients with locally advanced nasopharyngeal carcinoma [26]. Based on these promising results, there is a great interest in using early changes in tumor ADC values to adapt treatment plans based on early response, a process that can potentially be facilitated by high-frequency imaging on the MR-Linac. However, robust validation of DWI sequences on the MR-Linac is necessary, and to our knowledge, no studies in head and neck cancer have included DWI at a regular interval during RT using a MR-Linac device.

In this study, we aim to characterize the serial quantitative DWI changes in primary tumor and nodal target volumes in a pilot dataset of patients with HNSCC treated using the MR-Linac. According to the R-IDEAL framework, the study is considered a Stage 2a (“technical optimization of the innovation for treatment delivery”) [12]. This work represents a unique opportunity to evaluate DWIs obtained throughout the entire RT course, to quantify ADC changes on a weekly basis, and to correlate these changes with response to RT and subsequent oncologic and survival outcomes.

## MATERIALS AND METHODS

### Study Population

Patients with HNSCC who received treatment with curative-intent IMRT using a 1.5T MR-Linac (Elekta Unity; Stockholm, Sweden) at The University of Texas MD Anderson Cancer Center, from May 16^th^, 2019 to February 22^nd^, 2021 were included in this study.

The study was approved by the Institutional Review Board (Protocol number: PA18-0341) at MD Anderson Cancer Center and patients provided informed consent. The included patients are part of The Multi-OutcoMe EvaluatioN of radiation Therapy Using the Unity MR-Linac Study (MOMENTUM), which is a multi-institutional, observational international registry for patients treated on the MR-Linac system (NCT04075305).

Inclusion criteria were being at least 18 years old, having histologically confirmed HNSCC, having good performance status (Eastern Cooperative Oncology Group score of 0-2), receiving definitive RT for non-metastatic tumor, and having no contraindications for MRI [27].

Patients’ demographic data, disease characteristics, radiation therapy data and oncologic outcome were collected from the University of Texas MD Anderson Cancer Center clinical databases through the EPIC electronic medical record system by a manual review of clinical notes and paperwork.

### MR images

MRIs were acquired using the Unity system (Elekta AB, Stockholm, Sweden), which includes a Philips 1.5 T Marlin MRI with a 4-element anterior coil and a built-in 4 element posterior coil covering the head and neck region. MRIs were obtained for all patients at baseline (pre-RT) and on a weekly basis throughout the RT course. The following images were collected:

1. Three-dimensional (3D) T2-weighted MRI, which was used for image registration during daily treatment (repetition time = 1535 ms, echo time = 278 ms, pixel bandwidth = 740 Hz, flip angle = 90°, echo train length = 114, field of view = 400×400×300 mm^3^, reconstructed voxel size = 0.83×0.83×1 mm^3^, scan time = 2 minutes, number of average = 1, and SENSE factor = 4) (172 scans)
2. 3D T2-weighted MRI without fat suppression, which was used for target segmentation (repetition time = 2100 ms, echo time = 375 ms, pixel bandwidth = 459 Hz, flip angle = 90°, echo train length = 150, field of view = 520×520×300 mm^3^, and reconstructed voxel size = 0.98×0.98×2.2 mm^3^, scan time = 6 minutes, number of average = 2, and SENSE factor = 2) (9 scans)
2. 3D T2-weighted MRI with fat suppression, which was used for target segmentation (repetition time = 1400 ms, echo time = 190 ms, pixel bandwidth = 473 Hz, flip angle = 90°, echo train length = 76, field of view = 520×520×300 mm^3^, and reconstructed voxel size = 0.98×0.98×1.2 mm^3^, scan time = 6 minutes, fat saturation = SPAIR, number of average = 2, and SENSE factor = 2) (1 scan)
3. Single-shot echo planar DWI covering targets and organs-at-risk, which was used for treatment assessment (b values = 0, 150, and 500 s/mm^2^; repetition time = 5700 ms; echo time = 75 ms; pixel bandwidth = 2174 Hz; flip angle = 90°; echo train length = 39; field of view = 300×300×158 mm^2^; reconstructed voxel size = 1.6×1.6×1.3 mm^3^; scan time = 3 minutes; fat saturation = SPAIR; and SENSE factor = 2.2).
4. ADC maps were reconstructed using only the b-values of 150 and 500 s/mm^2^; the b=0 images were excluded to minimize the effects of perfusion on ADC calculations and to be consistent with MR-Linac Consortium recommendations [10]. These ADC maps were subsequently used to extract the histogram parameters of the segmented regions of interest (ROIs).

### Image Segmentation and Registration

Manual segmentation of the different ROIs, including the gross primary disease volume (GTV-P) and gross nodal disease volumes (GTV-N) was performed by two expert radiation oncologists: DE and ASRM with 9 and 15 years of experience, respectively. The baseline primary disease volume, manually segmented on the baseline T2-weighted images, was labeled as GTVP-BL. Then the ROIs were propagated to the rigidly co-registered DWIs. Subsequently, deformable image registration was performed to co-register the images of different weeks with the baseline images. The baseline primary tumor volumes were then propagated from the baseline images to the co-registered weekly time points. For each weekly image, the residual primary disease volume was also manually segmented using the T2-weighted images and labeled as GTVP-RD. The response sub volumes were created by subtracting the propagated initial GTVP-BL at each week minus the GTV-RD and was labeled as GTVP-RS. Propagation of all ROIs, from the T2-weighted images to the co-registered DWI of the same time point was done for all time points as illustrated in Figure. 1. The process of manual segmentation and registration was performed using the benchmarked commercially available image registration software Velocity AI (version 3.0.1). Finally, ADC measurements were extracted to assess ADC changes of the target volumes on a weekly basis relative to the baseline.

**Figure 1.**
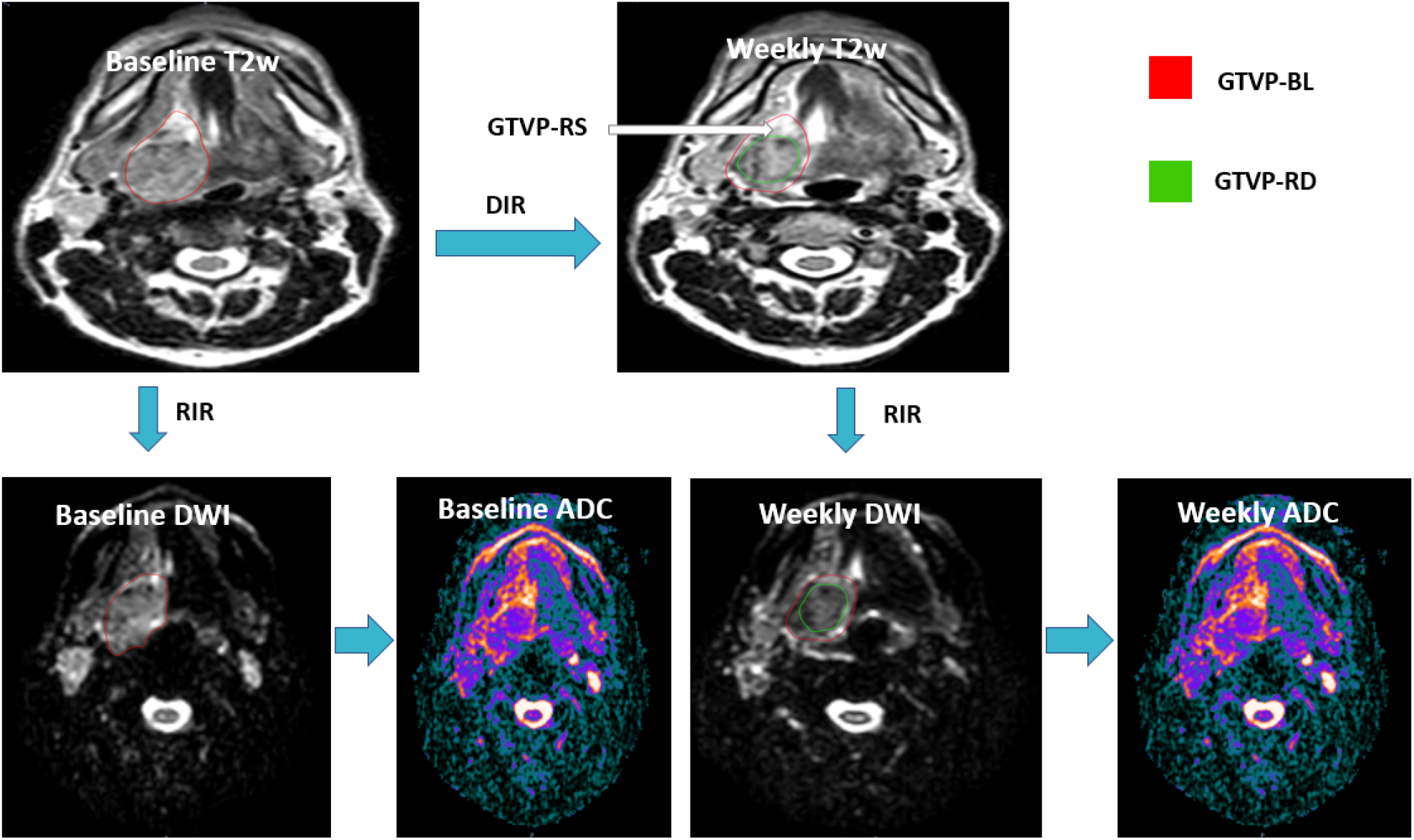
Image segmentation and registration workflow T2w: T2 weighted DWI: Diffusion weighted image ADC: Apparent diffusion coefficient RIR: Rigid image registration DIR: Deformable image registration GTVP_BL: Baseline primary tumor volume GTVP_RD: The residual primary disease volume GTVP_RS: The volume of the primary disease responding to RT

### Follow Up and Clinical Outcomes

All the patients underwent physical examination, fiberoptic endoscopy, contrast-enhanced computerized tomography, fluorodeoxyglucose–positron emission tomography CT and MRI, performed 8 to 12 weeks after the end of radiation therapy for treatment response assessment. Response to treatment was defined as occurrence of complete remission (CR) or not based on the Response Evaluation Criteria in Solid Tumors, version 1.1. Time to CR, local control, recurrence free survival (RFS), and overall survival (OS) were the calculated endpoints.

### Statistical analysis

The collected data were tabulated and analyzed using JMP pro, version 15. Continuous data were described as mean ± standard deviation (SD) and categorical data as proportions. The ADC values for all voxels of different ROIs (i.e., GTV-P and GTV-N) were assessed by histogram analysis. The following ADC histogram parameters were extracted using an in-house MATLAB script (MATLAB, MathWorks, MA, USA): mean, 5^th^, 10^th^, 20^th^, 30^th^, 40^th^, 50^th^ (i.e., median), 60^th^, 70^th^, 80^th^, 90^th^, and 95^th^ percentiles. We examined the relationship between these ADC parameters (at the baseline and weekly during RT) versus treatment response (CR vs. non-CR)–during and after the RT course–and the development of recurrence, using the non-parametric Mann-Whitney U test. The non-parametric Wilcoxon signed-rank test was used to compare the weekly ADC histogram parameters to the baseline. We used Bonferroni correction for multiple comparison across different weeks and P < 0.008 was considered significant. ΔADC, defined as the percent change of the ADC histogram parameter at each week (Wx) of the RT course relative to the baseline value for each ADC parameter, was calculated using the following equation: 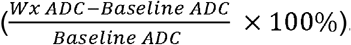. Also, weekly volumetric changes for both

GTV-P and GTV-N at weekly RT were calculated, and the non-parametric Spearman rho test was used to determine the strength and direction of the relationship between ΔADC and change in volume. Recursive partitioning analysis (RPA) was performed to identify ΔADC threshold associated with different oncological outcomes, such as the development of CR and relapse.

Additionally, we analyzed the kinetics of ADC changes for the HPV+ versus HPV unrelated head and neck cancer patients to demonstrate the specific profile of ADC changes in each cohort.

Missing ADC parameter values (due to a few patients with missing weekly images) were imputed based on the remaining timepoints available for a given patient using an order-1 linear spline method performed using an in-house Python script (Python version 3.8.8).

In this work, we followed the checklist of items to be reported per the REporting recommendations for tumor MARKer (REMARK) guidelines for prognostic studies [28].

## RESULTS

### Patient and Disease Characteristics

Thirty patients with HNSCC were included in this pilot study; most patients were male (n=28, 93.3%). At baseline, 18 patients had both primary tumor and lymph node involvement. Nine patients exhibited a primary tumor without lymph node involvement, and 3 patients had nodal disease only. Those who did not have primary tumor had either carcinoma of unknown primary or had their primary tumor surgically resected (i.e., tonsillectomy). Oropharyngeal carcinoma represented the most common type of HNSCC (n=21, 70%), followed by laryngeal cancer (n=7, 23.3%). Twenty-two patients (73.3%) had positive test results for human papilloma virus, with 20 patients having oropharyngeal primary tumors, 1 patient having carcinoma of unknown primary, and 1 patient having laryngeal carcinoma. Twenty-five patients (83.3%) had early-stage tumors (stage I and II), as classified by the American Joint Committee on Cancer guidelines, 8th edition. A summary of the patients’ and disease characteristics are shown in Table 1.

**Table 1.** Patients’ and Disease Characteristics:

| **Patient characteristics** | **Mean ± SD, range** |
| --- | --- |
| **Age (years)** | 64.1 ± 10.37, 37-82 |
| **Gender** |  |
| Male | 28 (93.3%) |
| Female | 2 (6.7%) |
| **Anatomical location** |  |
| Oropharynx | 21 (70%) |
| Larynx | 7 (23.3%) |
| Hypopharynx | 1 (3.3%) |
| CUP | 1 (3.3%) |
| **TNM Stage** |  |
| I | 21 (70%) |
| II | 4 (13.3%) |
| III | 3 (10%) |
| IV | 2 (6.7%) |
| **T stage** |  |
| Tx | 1 (3.3%) |
| T1 | 13 (43.3%) |
| T2 | 13 (43.3%) |
| T3 | 1 (3.3%) |
| T4 | 2 (6.7%) |
| **N stage** |  |
| N0 | 9 (30%) |
| N1 | 14 (46.7%) |
| N2 | 6 (20%) |
| N3 | 1 (3.3%) |
| **HPV status** |  |
| Positive | 22 (73.3%) |
| Negative | 1 (3.3%) |
| unknown | 7 (23.3%) |
| **Smoking Status** |  |
| Smoker | 1 (3.3%) |
| Non-smoker | 15 (50%) |
| Ex-smoker | 14 (46.7%) |
| **Surgery for the primary** |  |
| Yes | 7 (23.3%) |
| No | 23 (76.7%) |
| **RT dose (Gy)** | 68.5 ± 2.45, 63-70 |
| **Number of fractions** | 32.2 ± 1.93, 28-35 |
| **Chemotherapy** | 19 (63.3%) |
| Induction CT | 1 (3.3%) |
| Concurrent CT | 18 (60%) |
| **CR for the primary during RT course** |  |
| Yes | 11 (36.7%) |
| No | 19 (63.3%) |
| **CR at the end of RT** |  |
| Yes | 26 (86.7%) |
| No | 4 (13.3%) |
| **Recurrence** |  |
| Yes | 5 (16.7%) |
| No | 25 (83.3%) |
| **Type of recurrence** |  |
| Local | 1 |
| Regional | 0 |
| Distant | 4 |

### Oncologic Outcomes

The median follow-up time for the patients in this study was 19.6 months (range, 11.5-39.8 months), with 2-year local control, RFS and OS rates of 86. 7%, 83.3%, and 96.7%, respectively. Throughout the RT course, 11 of the 27 patients who had GTV-P had a CR for the primary tumor. Of these 11 patients, 5 patients had a CR at the 6th week of RT, 3 at the 5th week, 2 at the 4th week, and 1 at the 3rd week. Twenty-six patients (86.7%) had a complete resolution of both the primary tumor and the metastatic lymph nodes when evaluated 8 to 12 weeks after the end of RT. No patients had a CR of the lymph node metastases during RT. During surveillance, recurrence occurred for 5 patients (16.6%), with only one having local recurrence and the remaining 4 patients having distant recurrence. All patients had their recurrent disease pathologically confirmed.

### ADC Kinetics in Relation to treatment Response and Oncologic Outcome

All studied pre-treatment ADC parameters did not have a significant association with the response to RT or the oncologic outcomes (p > 0.05).

Overall, there was a significant increase, starting from the 2^nd^ week of RT, in propagated initial GTV-P mean ADC, compared to baseline mean ADC (p = 0.015, 0.002, <0.001, 0.001 and 0.007, respectively). However, this increase in mean ADC reached statistical significance only for primary tumors developing CR during RT. Similarly, a statistically significant incremental increase in GTV-N mean ADC compared to the baseline mean ADC (p < 0.001 and 0.005, respectively) was found in all patients (Figures. 2 and 3).

**Figure 2.**
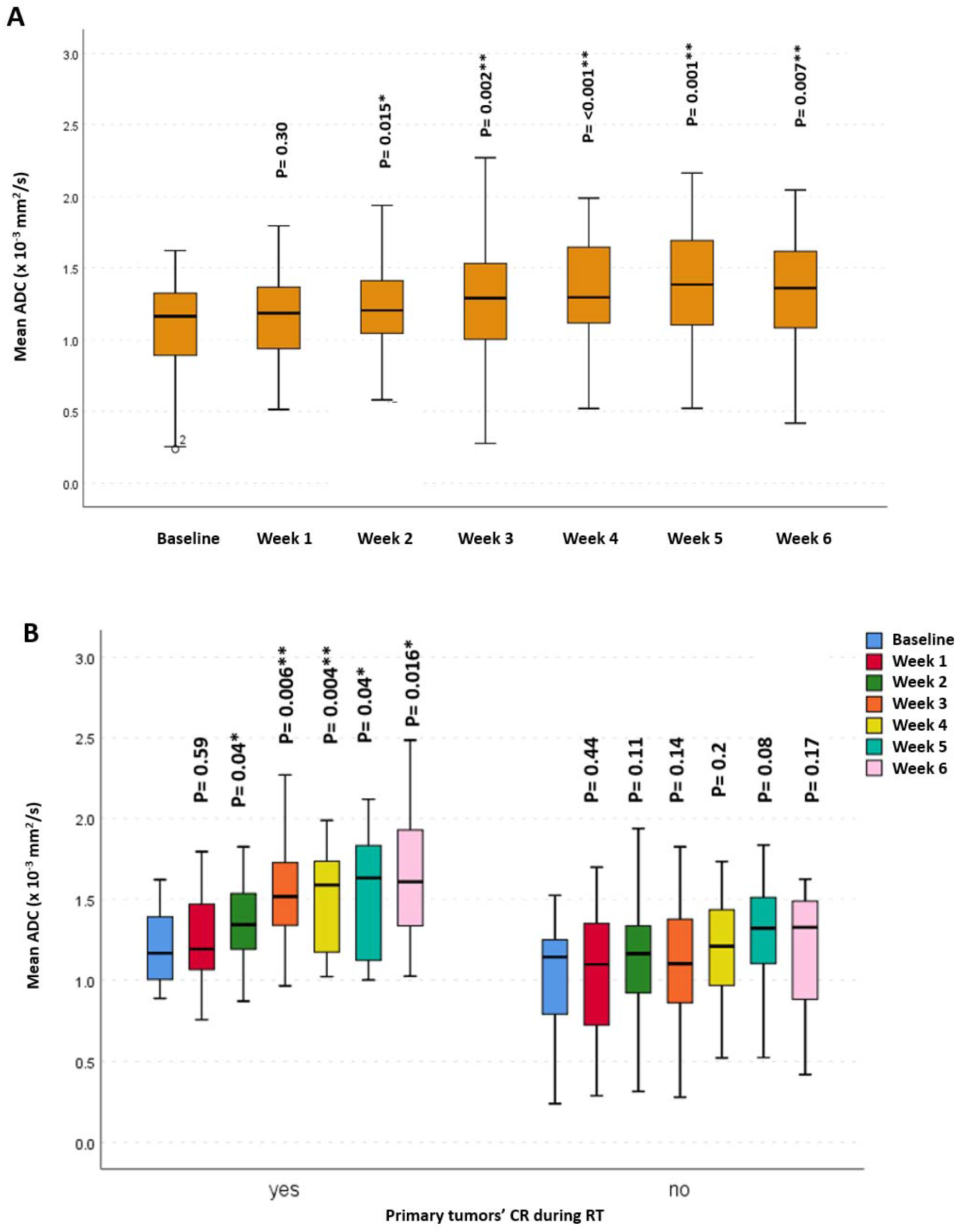
Mean apparent diffusion coefficient (ADC) at different timepoints for GTV-P: (A) For all patients, (B) patients who developed CR during RT versus those who did not.

**Figure 3.**
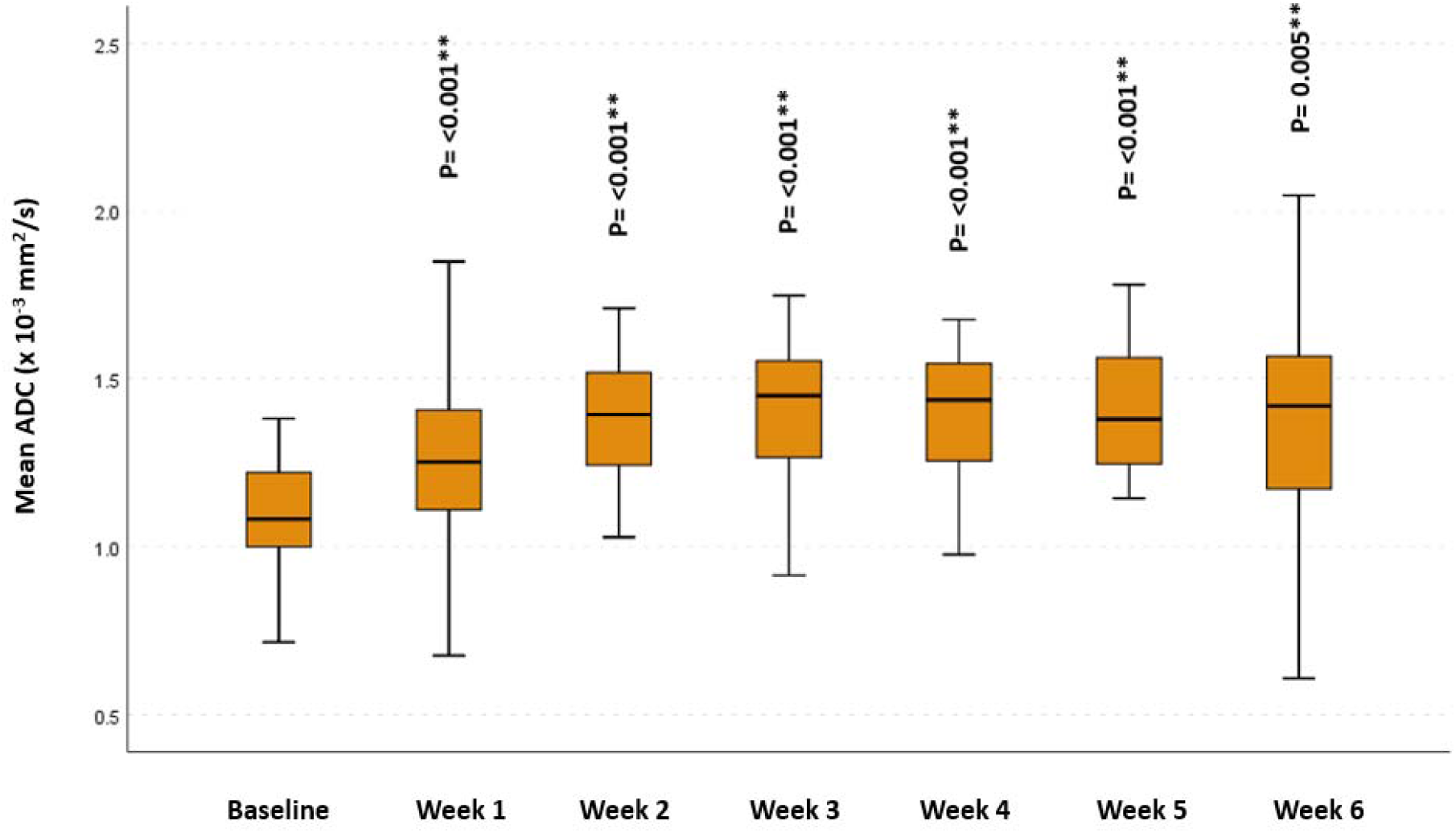
Mean ADC at different timepoints for GTV-N *Significance before Bonferroni correction ** Significance after Bonferroni correction

All other ADC parameters for the propagated initial GTV-P showed a significant increase from the baseline throughout the course of RT (Figure. 4).

**Figure 4.**
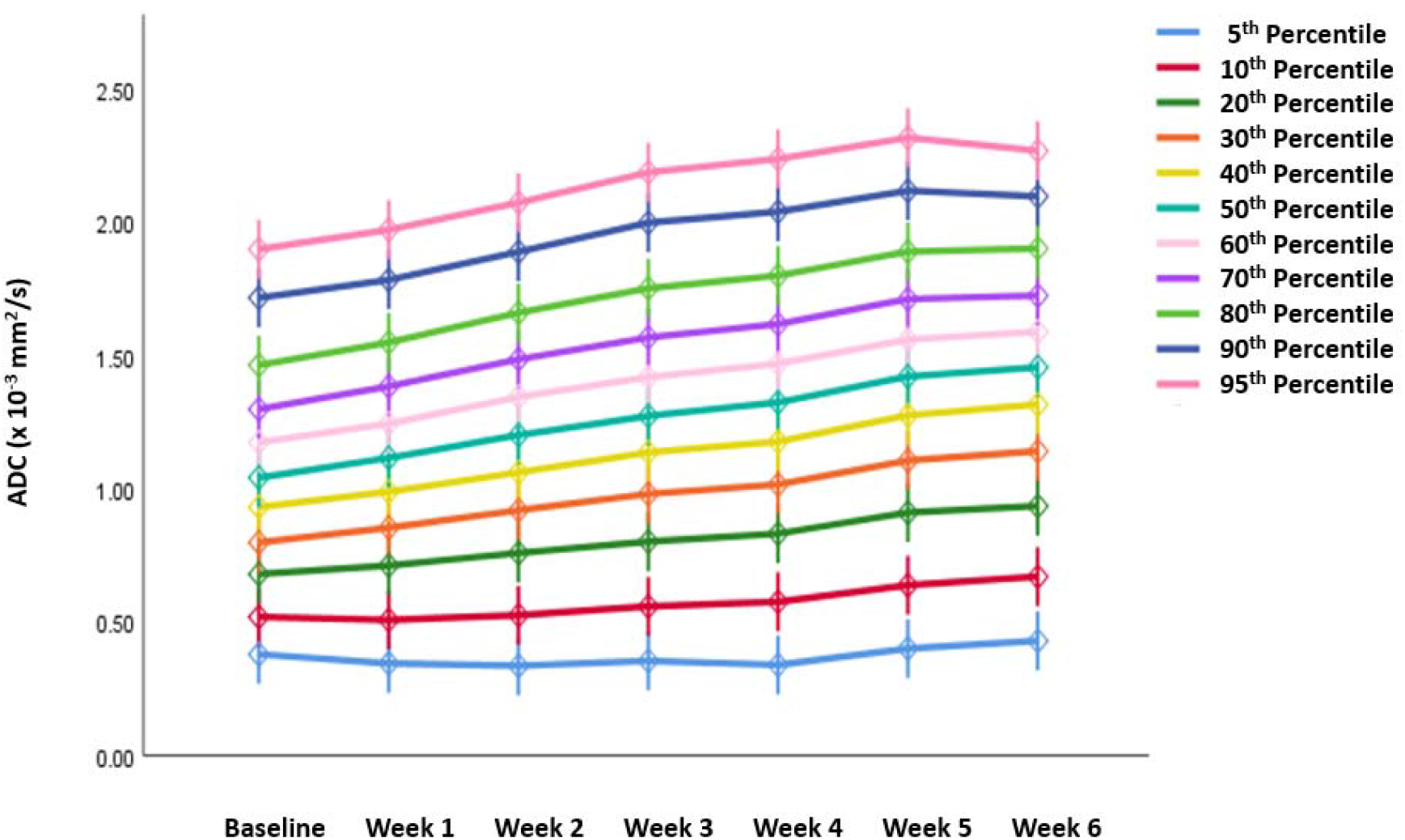
Absolute ADC histogram parameters for GTV-P across different time points.

A significant association was detected between ΔADC 5^th^, 10^th^, 20^th^, 30^th^, 40^th^, 60^th^ and 70^th^ percentiles at the 3^rd^ week of RT for propagated baseline GTV-P, and occurrence of CR for the primary tumor, during RT (n=11, 40.7%) (p = 0.029, 0.036, 0.042, 0.049, 0.048, 0.034, and 0.048, respectively). RPA identified a ΔADC 5^th^ percentile > 13% at the 3rd week of RT as the most significant parameter associated with shorter time to CR for the primary tumor during RT (p < 0.001). However, there was no statistically significant correlation between GTV-P ΔADC parameters at various time points and development of CR after the end of RT (n = 26, 86.7%). Additionally, there was no significant association between ΔADC parameters and RFS.

No significant correlation was found between mean ΔADC for GTV-N and other oncologic outcomes.

### Analysis of Volumetric and Mean ΔADC Changes

There was a significant decrease in residual tumor volumes for both GTV-P and GTV-N compared to baseline volumes at different time points throughout the RT course. Although this gradual decrease in the residual primary tumor’s volume was detected earlier, starting from the 1^st^ week of RT, the GTV-N volumetric decline was first demonstrated at week 3 (Figure. 5).

**Figure 5.**
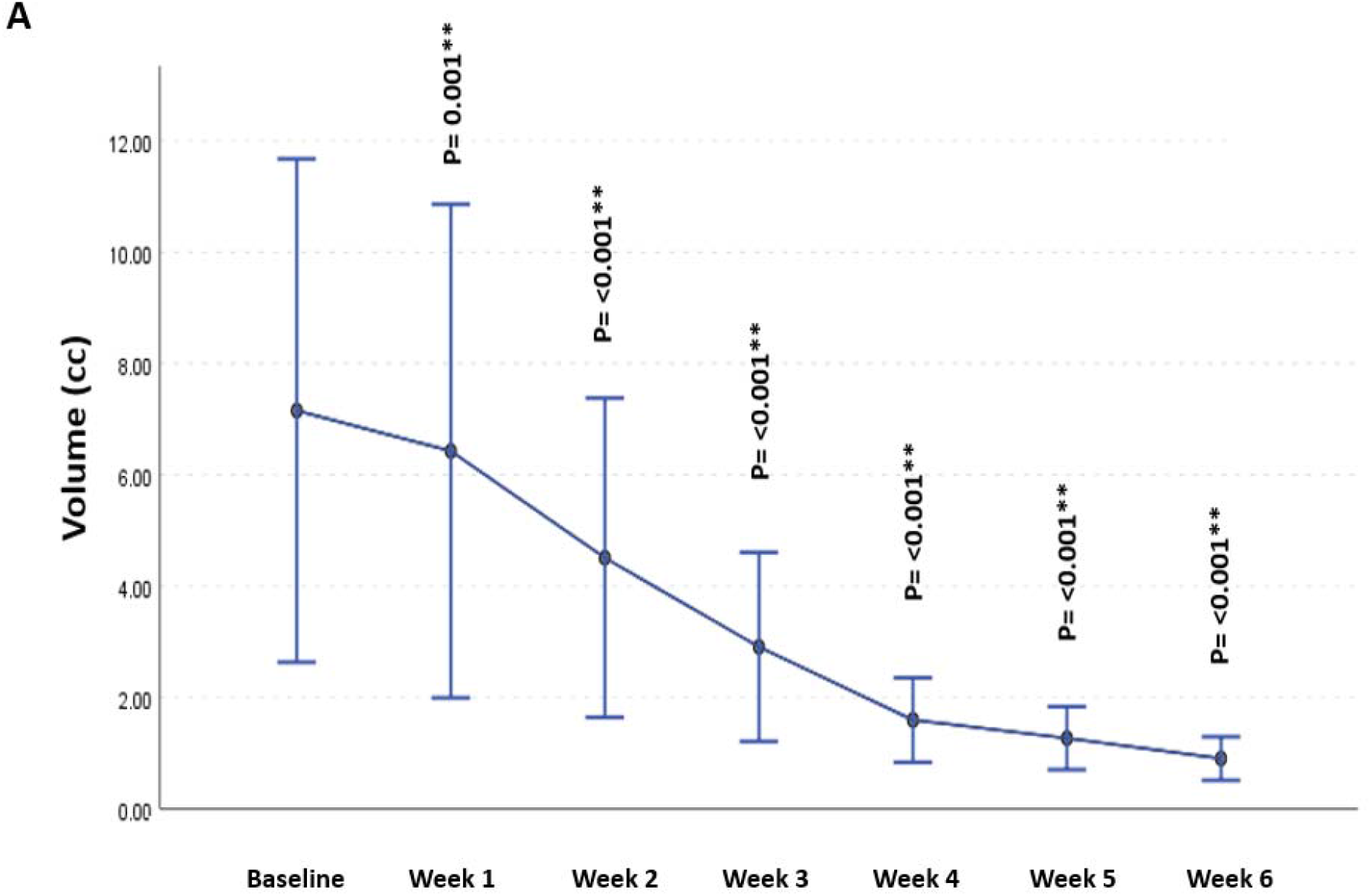
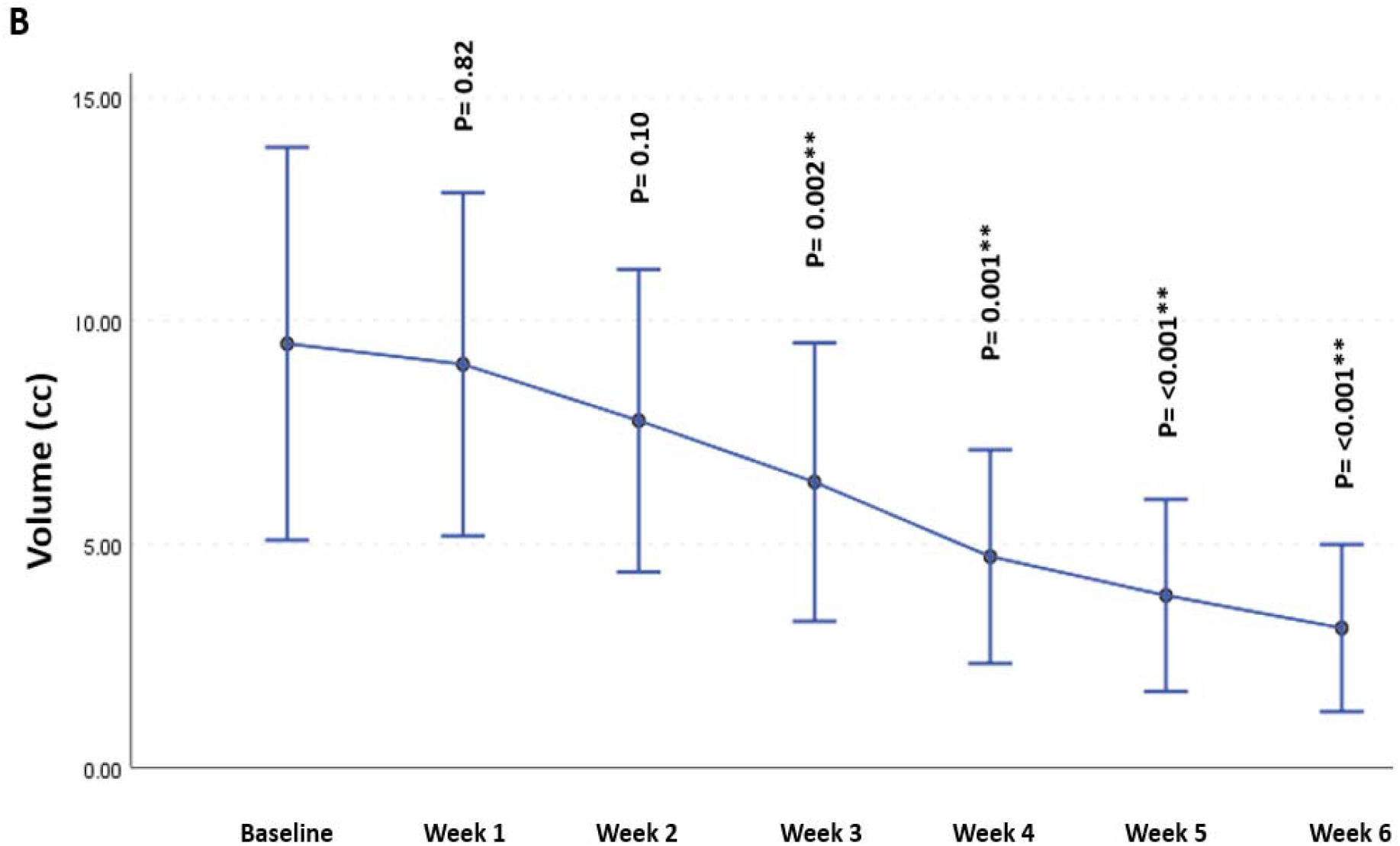
Volumetric changes in GTVP-RD (A) & GTV-N (B) throughout the course of radiation therapy *Significance before Bonferroni correction ** Significance after Bonferroni correction

A statistically significant negative correlation was demonstrated between the mean ΔADC and change in volume for GTVP-RD at the 3rd and 4th weeks of RT (rho = -0.39, p = 0.044, and rho =-0.45, p =0.019, respectively; Figure. 6). However, no significant correlation was found between mean ΔADC and change in volume for GTV-N.

**Figure 6.**
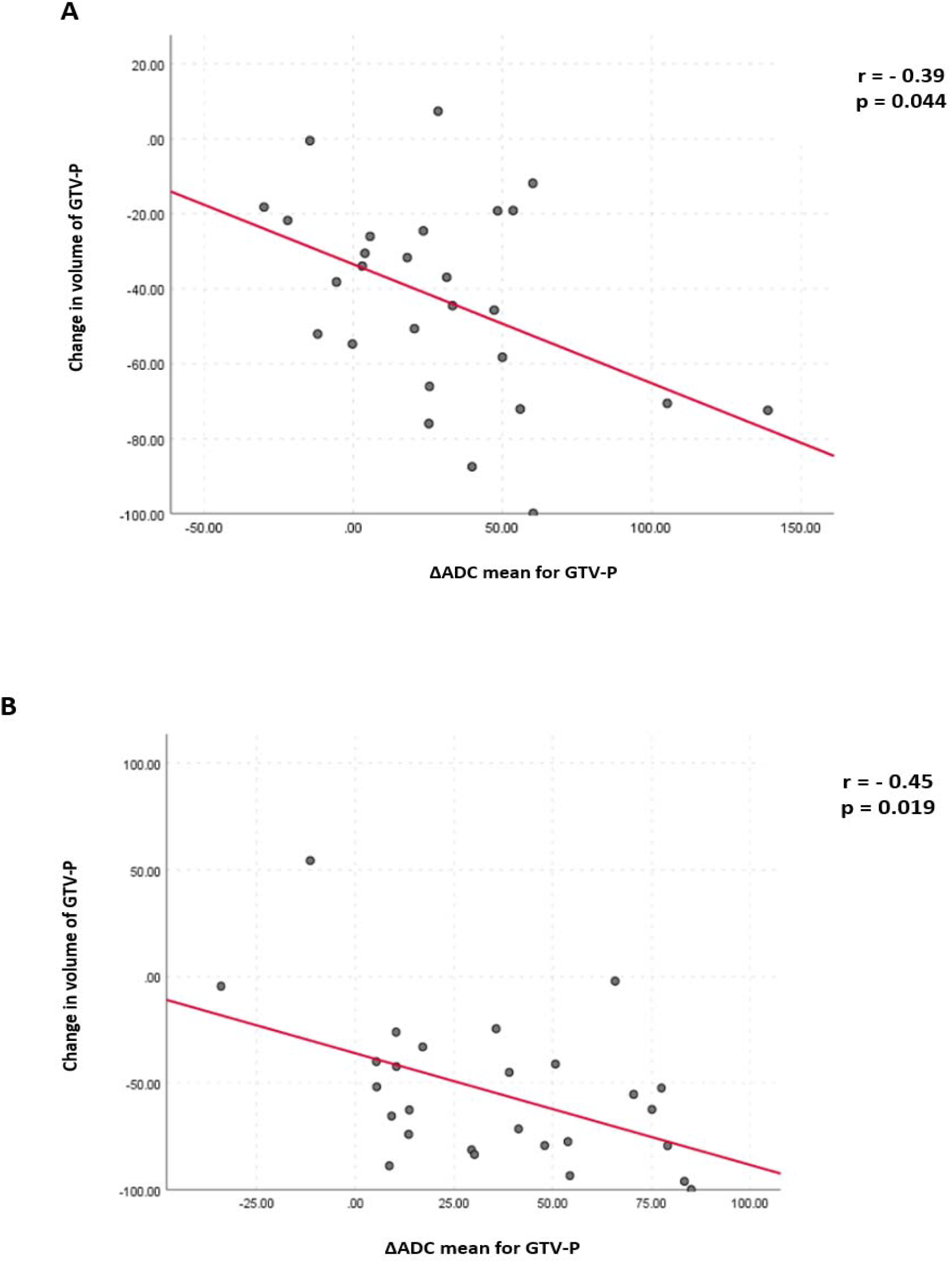
The relationship between changes in volume and mean ADC for GTVP-RD: (A) at the 3^rd^ week (B) at the 4^th^ week of radiation therapy

There was no significant correlation (p > 0.05) between volumetric changes, either in GTV-P or GTV-N, and the different oncologic outcomes at the end of RT (CR or recurrence).

### ROI Sub-Volume Analysis

As detected for the propagated initial GTV-P, there was an incremental increase in GTVP-RD mean ADC compared to the baseline values. This rise in GTVP-RD mean ADC was first demonstrated at the 2^nd^ week of RT.

A significant correlation was detected between mean ΔADC at the 3^rd^ week of RT for GTVP-RD and the occurrence of CR for the primary tumor, during RT (p = 0.043). RPA identified a mean ΔADC > 20% at the 3rd week of RT as the most significant parameter associated with shorter time to CR (p = 0.013).

### HPV+ Versus HPV Unrelated subgroup analyses

We had 22 patients with HPV+ and 8 patients with HPV unrelated head and neck cancer patients included in this study. For HPV+ cases, 11 patients had their primary tumor originating from tonsils, 9 from base of tongue, one patient had his primary tumor not identified (carcinoma of unknown primary), and one patient had laryngeal tumor. On the other hand, larynx (n = 6, 75%) was the most common site harboring primary tumor followed by both oropharynx and hypopharynx (one for each) in HPV unrelated patients. In HPV+ group, half of the primary tumors completely resolved during RT, while none of the primary tumors in the HPV unrelated group achieved CR during treatment.

In HPV+ group, there was a significant increase in mean ADC (for both GTV-P & GTV-N) compared to baseline mean ADC. This incremental increase in the GTV-P mean ADC was statistically significant only for primary tumors developing CR intra-treatment (n = 11, 50%). In contrast, no significant changes in the GTV-P mean ADC was detected compared to the baseline values, in the HPV unrelated group.

For the HPV+ group, RPA identified a ΔADC 5^th^ percentile > 13% at the 3rd week of RT as the most significant parameter associated with shorter time to CR for the primary tumor during RT (p = 0.001).

For both groups, no statistically significant correlation was found between GTV-P or GTV-N ΔADC parameters at various timepoints and development of either CR (n = 18, 81.8% for HPV+ and n = 8, 100% for HPV unrelated) or recurrence (n = 3, 13.6% for HPV+ and n = 2, 25% for HPV unrelated) after the end of RT.

There was a significant decrease in residual tumor volumes for both GTV-P and GTV-N compared to baseline volumes at different timepoints throughout the RT course, in the HPV+ group.

Results for HPV+ and HPV unrelated subgroup analyses is shown in supplement 1.

## DISCUSSION

In this prospective study, quantitative DWI obtained weekly throughout the RT course were used to determine the gross tumor diseases’ ADC kinetics, in a granular form during RT. These ADC changes were correlated with response during and after RT and oncologic outcomes.

Our study demonstrated a statistically significant rise in ADC parameters during RT compared to baseline values in all patients. The increased mean ADC values, starting from the 2^nd^ week during RT, were found to be higher in patients who developed radiologic CR for the primary tumor during RT. These findings match the findings of previous studies highlighting the correlation between ADC changes and response to RT in HNC. For example, in a study conducted by Hatakenaka et al a higher ADC increase early during RT compared to baseline revealed significant differences between local control and failure of the primary tumors in patients with HNSCC [29]. Moreover, a prospective study, including 37 patients, has shown the significant intra-treatment increase in mean ADC (p < 0.001) for patients with local control compared to those with local failure [20]. The exact timing of these ADC changes was not clear in the previous publications. One of the most important findings in our study is that GTV-P mean ADC for patients with a favorable response to RT started to significantly increase in the 2nd week of treatment and continued until the end of RT. Additionally, in our pilot data, the most significant correlation between the primary tumors’ ΔADC at different parameters and CR of the primary tumor during RT appears to be noticeable at the 3rd week of treatment. RPA has identified a 13% increase in GTV-P ΔADC 5^th^ percentile at the 3rd week relative to the baseline ADC 5^th^ percentile as the most significant factor for predicting CR for the primary tumor during RT. A recent study conducted by our group showed an increase of 7% in the GTV-P mean ΔADC at mid-RT, in comparison to the baseline, was significantly associated with better locoregional control [25]. Upon conducting a post-hoc analysis using mid-RT (3^rd^ and 4^th^ weeks of RT) mean ΔADC threshold of 7%, we found that the CR rate for primary tumors during RT was higher for cases where the mean ΔADC > 7%. However, statistical significance was not reached with p values of 0.09 and 0.053, respectively. The identification of tumors developing CR during RT might be better associated with local control and oncologic outcomes in head and neck cancer patients as suggested in previous studies. Jaulerry et al demonstrated that tumor regression during RT was an independent predictive factor of local control in their patient population [30]. Additionally, a prospective study involving 152 patients with oropharyngeal and hypopharyngeal cancers treated with IMRT reported the rate of tumoral reduction during RT to be predictive for local control in those patients [31]. Although a significant correlation between higher ΔADC parameters and the occurrence of CR for the primary tumor during RT was detected, no significant correlation was found between ΔADC and either the response post-treatment or recurrence. This is possibly due to the small number of patients who did not achieve CR after the end of RT or experienced recurrence at a later stage. These findings contradict the previous data showing a significant association between ΔADC and both locoregional control and RFS at the end of RT [32, 33]. These ADC changes were assessed for the GTV-P propagated unchanged from the baseline images to weekly MRIs after image registration. This means that the previously described ΔADC parameters were independent from the volumetric changes in the primary tumor.

In terms of primary residual tumor (GTVP-RD), RPA found a ΔADC mean more than 20%, at the 3^rd^ week of RT to be significantly correlated with a shorter time to CR for the primary tumor during RT.

Our data could not reach a conclusion regarding the association of serial ADC changes and endpoints because of the nature of this relatively small pilot study with a limited number of events. There was no significant association detected between the ADC parameters, extracted from GTV-N and loco-regional control or RFS. This finding might be explained by the absence of regional recurrence in our cohort. Moreover, the heterogeneity of the lymph node metastases (presence of a cystic and solid component) may contribute to the absence of correlation between the nodal ADC parameters and oncologic outcomes.

We also showed that baseline ADC parameters, whether in the primary tumor or lymph node metastases, had no significant correlation with different oncologic outcomes, indicating that dynamic information obtained from RT-induced imaging changes during treatment is likely more informative compared to baseline status. This finding is in agreement with previous studies by our group as well as by other groups [25, 34, 35]. However, these findings conflict with some studies that showed a significant association between pre-treatment ADC parameters and the response to treatment [29, 36].

The volumetric analysis for different ROIs, showed a significant decrease in the GTVP-RD volume during RT compared to the baseline volume. This volumetric decrease was observed immediately starting from the 1^st^ week of RT. However, a delayed decremental decline in GTV-N volumetric changes, starting from the 3^rd^ week of RT, was demonstrated. This finding may justify why lymph node metastases did not attain CR during RT in contrast to the higher rate of intra-treatment CR achieved by primary tumors.

Another important finding in our study, is the significant negative correlation that was found between mean ΔADC and volumetric changes of GTVP-RD, at the 3^rd^ and 4^th^ weeks of RT however, no significant correlation was detected between changes in both mean ADC and volume of GTV-N. This finding agrees with similar findings by Ng at al. [37]. It is noteworthy that our finding is consistent with the fundamental biological principles underlying DWIs, in which diffusion of water is dependent on cellular density of tissues; therefore, a reduction in tumor cellularity in response to RT would result in increases in ADC parameters.

When comparing HPV+ versus HPV unrelated tumors, the HPV+ group showed similar profile of imaging changes. The mean ADC for gross tumor disease (primary tumor and metastatic lymph nodes) showed a significant increase in HPV+ group compared to baseline. Only primary tumors completely resolved during RT showed a significant rise in mean ADC compared to the baseline. In contrast, no significant changes were observed in all the studied ADC histogram parameters at different timepoints when compared to the baseline ADC parameters in HPV unrelated cases. RPA identified a ΔADC 5^th^ percentile > 13% at the 3rd week of RT as the most significant parameter associated with shorter time to CR for the primary tumor during RT for HPV+ tumors. The number of patients developed either recurrence or non-CR post-RT was limited in both HPV+ and HPV unrelated groups, thus, no significant association was detected between various ADC parameters at different timepoints and oncologic outcomes. Furthermore, a significant reduction in the volume of both primary tumors and metastatic lymph nodes was observed in HPV+ cases compared to baseline volumes at different timepoints during RT.

This study has several limitations. First, a relatively small number of patients were included in the study. Additionally, a few patients had missing images due MR-Linac technical difficulties in image acquisition earlier at the time of initial implementation of the program.

Second, the limited number of locoregional failure or recurrence events resulted in the absence of meaningful correlation between different ADC parameters, throughout the course of RT, and clinical endpoints.

Furthermore, as ours was the first prospective study to investigate the role of quantitative MRIs obtained during treatment via MR-Linac, we intended to describe the kinetics of multiple ADC parameters at regular intervals and correlate these parameters with the overall outcome of patients with HNSCC. Therefore, our cohort constituted a heterogenous group of HNSCC with different primary sites of origin, representing another limitation of our study.

Finally, small primary tumors (i.e., T1 glottic carcinoma) and small size metastatic lymph nodes were included in the study. Small ROIs might have led to bias in measurement and extraction of ADC parameters, and consequently, bias in correlation between these ADC parameters and other endpoints.

## CONCLUSION

Our initial experience with serial DWI acquisition during RT using the MR-Linac showed that dynamic changes in ADC values, for both the primary tumor and nodal disease, assessed at regular intervals during RT can be a promising biomarker for predicting early response to treatment in head and neck cancer patients.

Further studies with larger cohorts of patients and more multi-institutional data, are needed to validate our results.

## Data Availability

All data will be available online at Figshare

doi:10.6084/m9.figshare.22766783

## Acknowledgements

We thank Ashli Nguyen-Villarreal, Associate Scientific Editor, and Bryan Tutt, Scientific Editor, in the Research Medical Library at The University of Texas MD Anderson Cancer Center, for editing this article.

