## Supplement for "Longitudinal diffusion and volumetric kinetics of head and neck cancer magnetic resonance on a 1.5T MR-Linear accelerator hybrid system: A prospective R-IDEAL Stage 2a imaging biomarker characterization/ pre-qualification study"

**HPV+ Vs HPV unrelated tumors**

**Table 1.1. HPV+ Patients’ and Disease Characteristics:**

| **Patient characteristics** | **Mean ± SD, range** |
| --- | --- |
| **Age (years)** | 65.43 ± 9.98, 37-81 |
| **Gender**  Male  Female | 21 (95.45%)  1 (4.54%) |
| **Primary sites**  Tonsil  BOT  CUP  Larynx | 11 (50%)  9 (40.9%)  1 (4.54%)  1 (4.54%) |
| **TNM Stage**  I  II  III  IV | 15 (68.2%)  2 (9%)  3 (13.6%)  2 (9%) |
| **T stage**  Tx  T1  T2  T3  T4 | 1 (4.54%)  7 (31.8%)  11 (50%)  1 (4.54%)  2 (9%) |
| **N stage**  N0  N1  N2  N3 | 2 (9%)  14 (63.63%)  5 (22.7%)  1 (4.54%) |
| **Smoking Status**  Non-smoker  Ex-smoker | 12 (54.54%)  10 (47.6%) |
| **Surgery for the primary**  Yes  No | 7 (31.8%)  15 (68.2%) |
| **RT dose (Gy)** | 69.55 ± 1.19, 66-70 |
| **Number of fractions** | 32.8 ± 1.03, 30-35 |
| **Treatment**  Radiation therapy alone  Concurrent Chemotherapy | 6 (27.3%)  16 (72.7%) |
| **CR for the primary during RT course**  Yes  No | 11 (50%)  11 (50%) |
| **CR at the end of RT**  Yes  No | 18 (81.8%)  4 (18.2%) |
| **Recurrence**  Yes  No | 3 (13.6%)  19 (86.36%) |
| **Type of recurrence**  Local and/ or regional  Distant | 0  3 |

**Table 2.1. HPV Unrelated Patients’ and Disease Characteristics:**

| **Patient characteristics** | **Mean ± SD, range** |
| --- | --- |
| **Age (years)** | 61/38 ± 11.83, 42-82 |
| **Gender**  Male  Female | 7 (87.5%)  1 (12.5%) |
| **Origin**  Larynx  Oropharynx  Hypopharynx | 6 (75%)  1 (12.5%)  1 (12.5%) |
| **TNM Stage**  I  II | 6 (75%)  2 (25%) |
| **T stage**  T1  T2 | 6 (75%)  2 (25%) |
| **N stage**  N0  N2 | 7 (87.5%)  1 (12.5%) |
| **Smoking Status**  Smoker  Non-smoker  Ex-smoker | 1 (12.5%)  3 (37.5%)  4 (50%) |
| **Surgery for the primary**  Yes  No | 0  8 (100%) |
| **RT dose (Gy)** | 65.67 ± 2.89, 63-69.96 |
| **Number of fractions** | 29 ± 2.41, 28-33 |
| **Treatment**  Radiation therapy alone  Induction Chemotherapy  Concurrent Chemotherapy | 5 (62.5%)  1 (12.5%)  2 (25%) |
| **CR for the primary during RT course**  Yes  No | 0  8 (100%) |
| **CR at the end of RT**  Yes  No | 8 (100%)  0 |
| **Recurrence**  Yes  No | 2 (25%)  6 (75%) |
| **Type of recurrence**  Local  Regional  Distant | 1 (12.5%)  0  1 (12.5%) |
| **Gross tumor disease**  GTV-P only  GTV-P & GTV-N  GTV-N only | 7 (87.5%)  8 (100%)  1 (12.5%) |

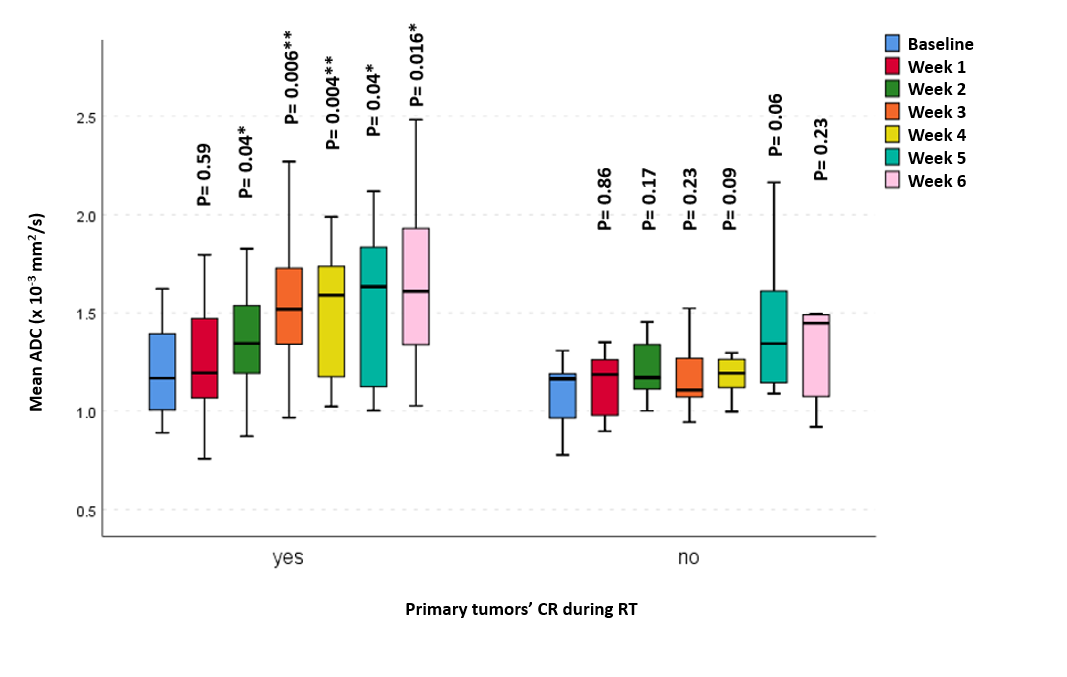

Figure 1.1 Mean apparent diffusion coefficient (ADC) at different timepoints for primary tumors which developed CR during RT versus those did not (HPV+).

*Significance before Bonferroni correction

** Significance after Bonferroni correction

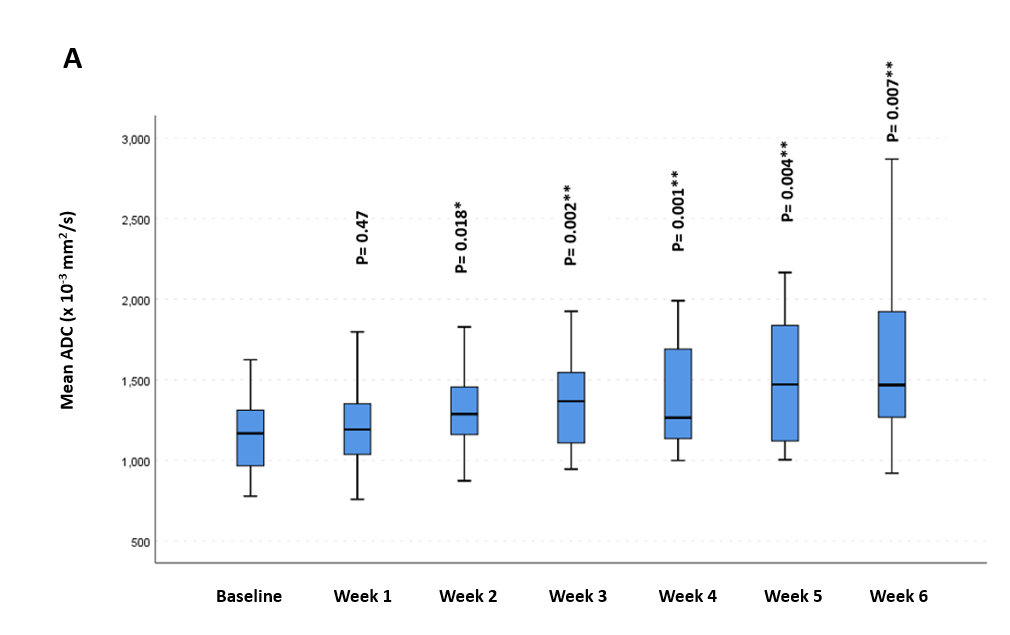

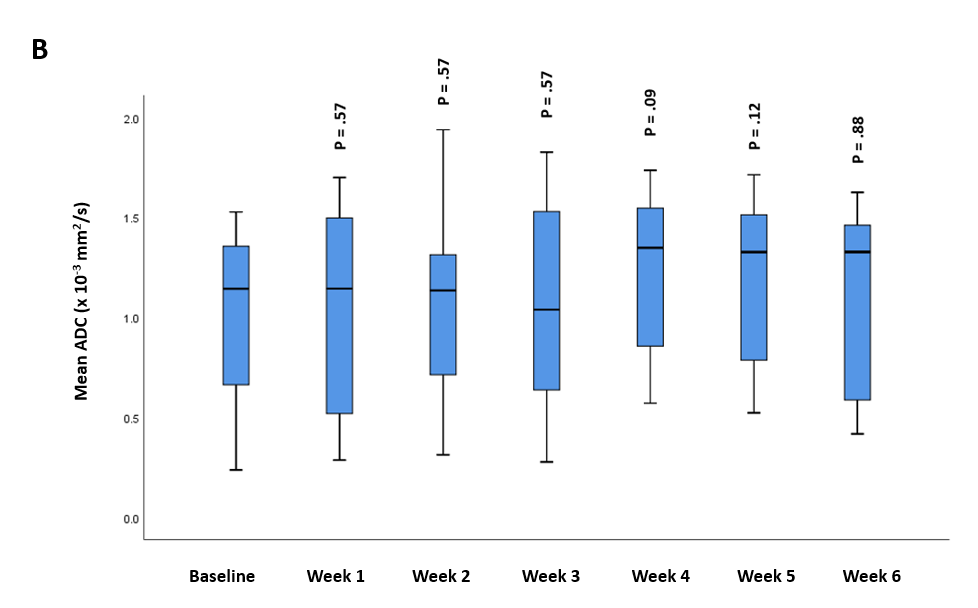

Figure 2.1. Mean ADC at different timepoints for GTV-P: (A) HPV+, (B) HPV unrelated

*Significance before Bonferroni correction

** Significance after Bonferroni correction

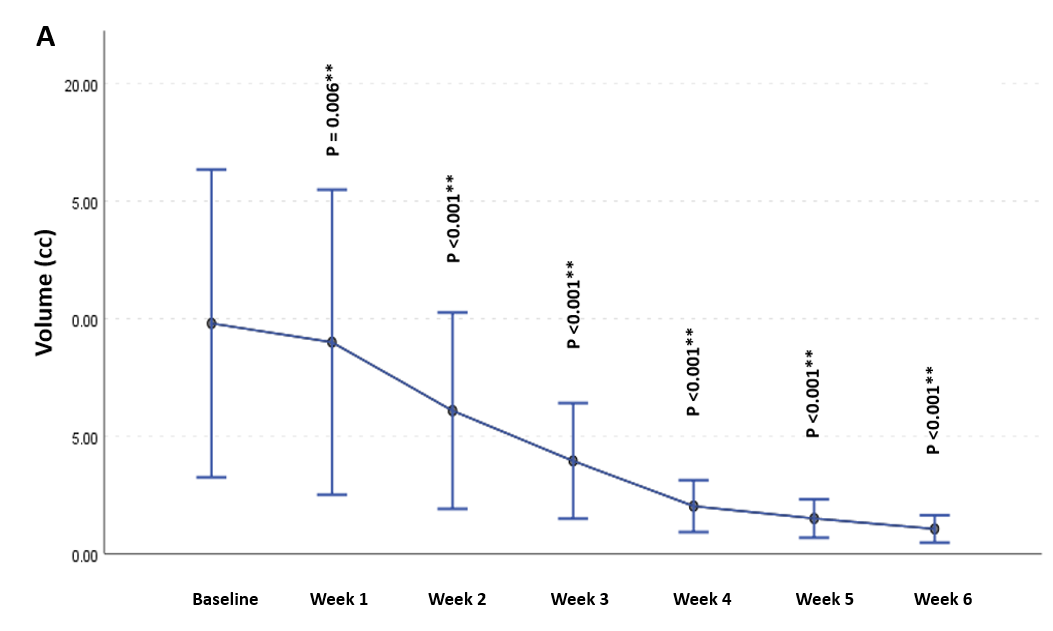

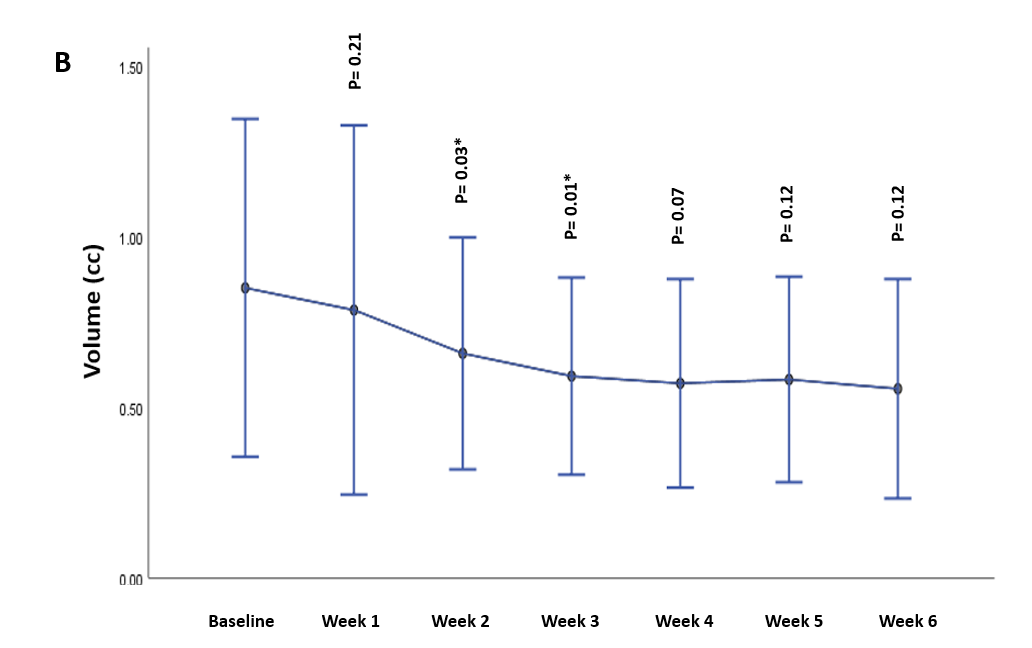

Figure 3.1. Volumetric changes in GTV-P throughout the course of radiation therapy: (A) HPV+ cases, (B) HPV unrelated

*Significance before Bonferroni correction

** Significance after Bonferroni correction

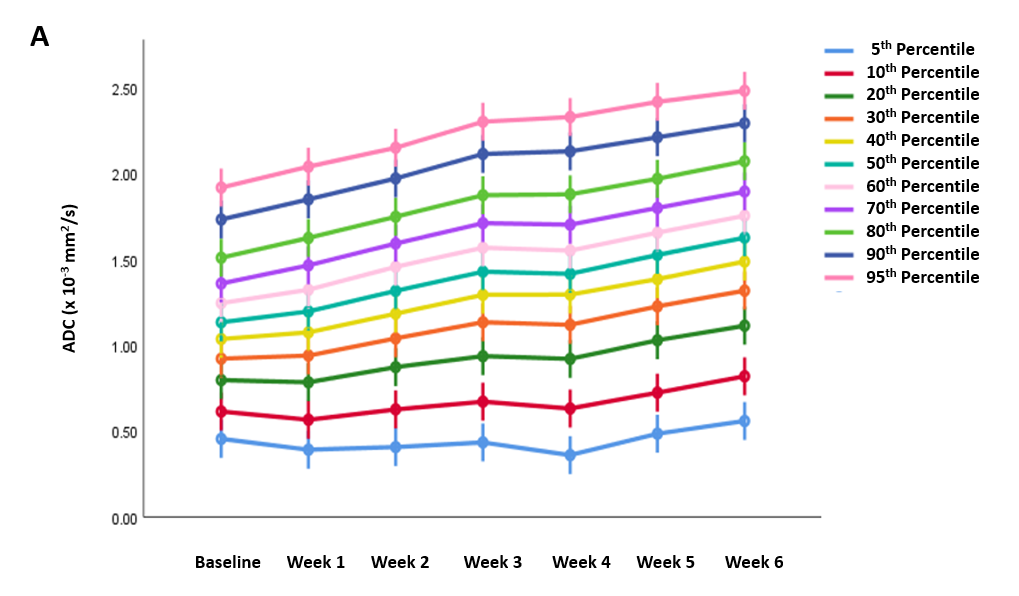

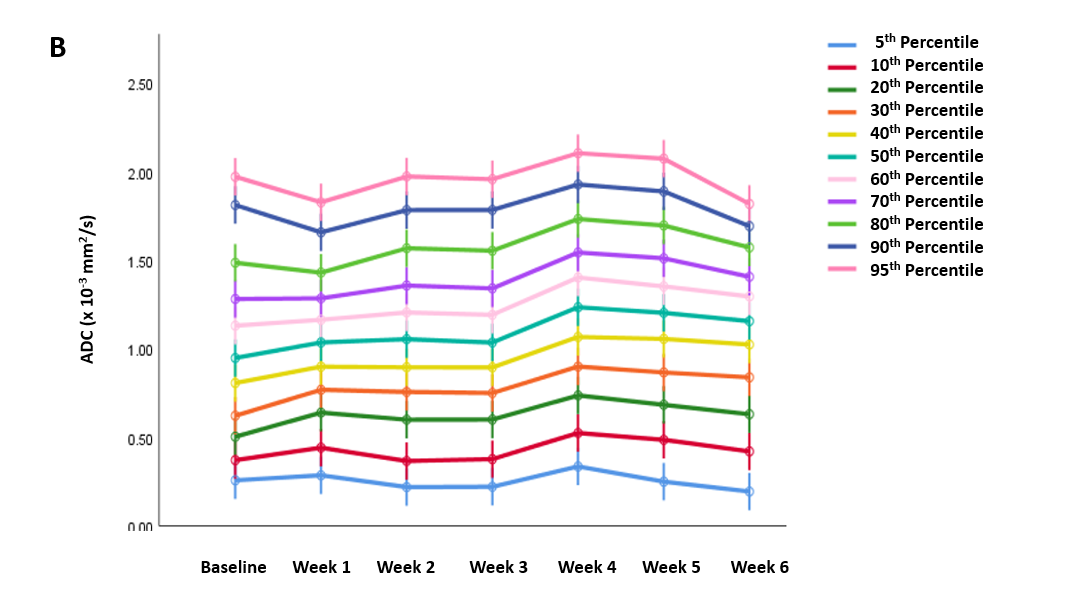

Figure 4.1. Absolute ADC histogram parameters for GTV-P across different time points: (A) HPV+, (B) HPV unrelated.

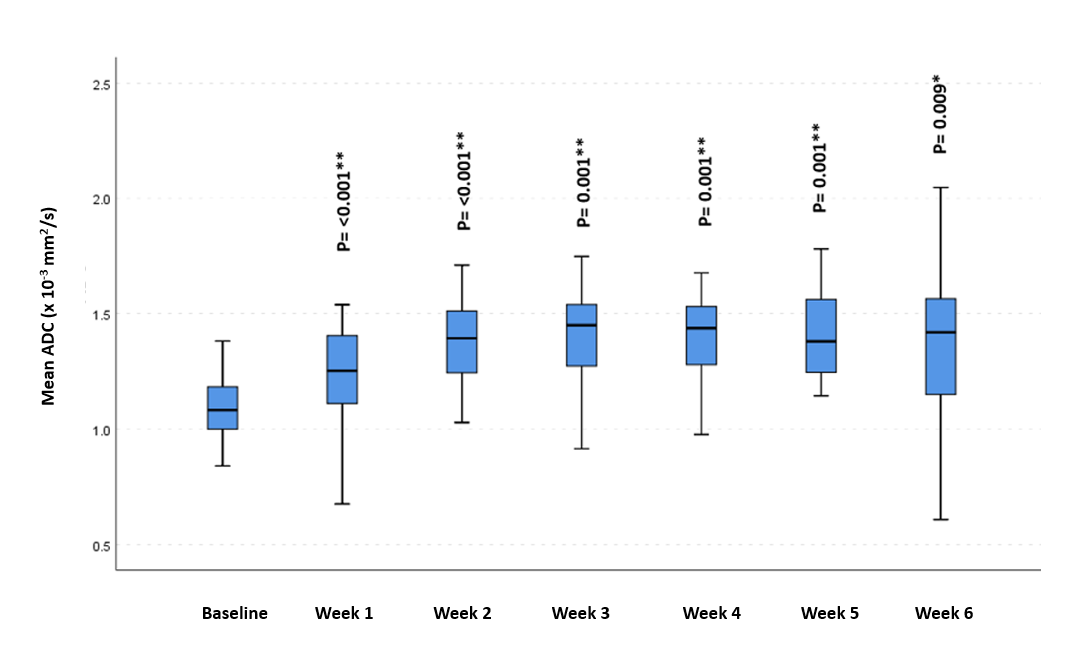

Figure 5.1. Mean ADC at different timepoints for GTV-N in HPV+ cases

*Significance before Bonferroni correction

** Significance after Bonferroni correction

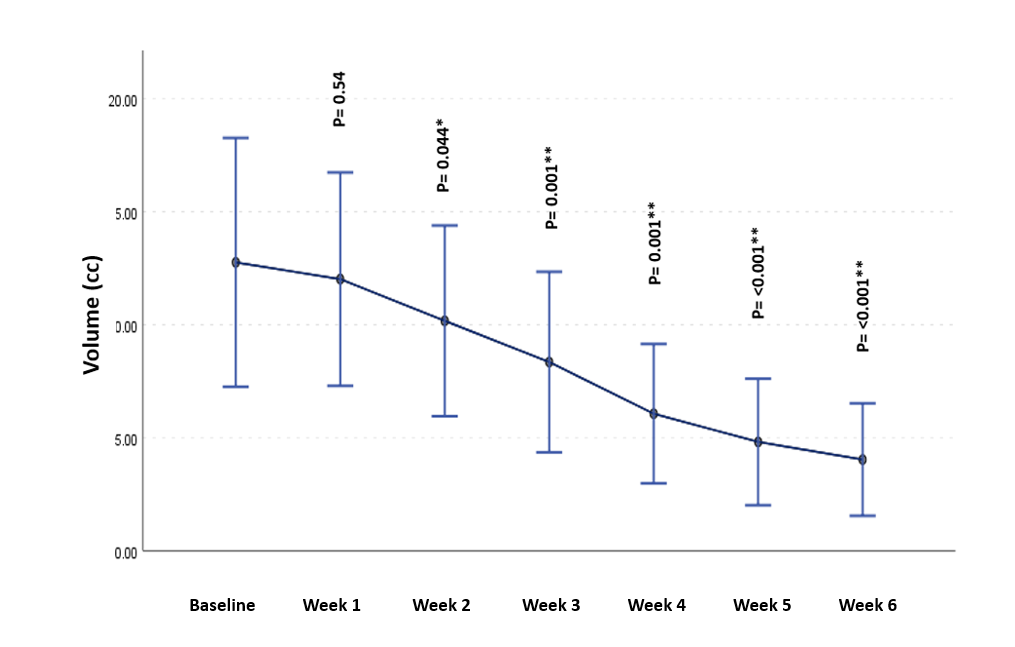

Figure 6.1. Volumetric changes in GTV-N throughout the course of radiation therapy in HPV+ cases

*Significance before Bonferroni correction

** Significance after Bonferroni correction
